# Comparing prognostic performance and reasoning between large language models and physicians

**DOI:** 10.64898/2026.04.17.26350898

**Authors:** Megan Gjertsen, WonJin Yoon, Majid Afshar, Brandon Temte, Brandon Leding, Stephen Halliday, Kaitlin Bradley, Joseph Kim, Juliana Mitchell, Anna K Sanders, Emma Croxford, John Caskey, Matthew Churpek, Anoop Mayampurath, Yanjun Gao, Timothy Miller, Jacqueline M Kruser

## Abstract

**Importance:** Physicians routinely prognosticate to guide care delivery and shared decision making, particularly when caring for patients with critical illnesses. Yet, these physician estimates are prone to inaccuracy and uncertainty. Artificial intelligence, including large language models (LLMs), show promise in supporting or improving this prognostication. However, the performance of contemporary LLMs in prognosticating for the heterogeneous population of critically ill patients remains poorly understood.

**Objective:** To characterize and compare the performance of LLMs and physicians when predicting 6-month mortality for hospitalized adults who survived critical illness.

**Design:** Embedded mixed methods study with elicitation and comparison of prognostic estimates and reasoning from LLMs and practicing physicians.

**Setting:** The publicly available, deidentified Medical Information Mart for Intensive Care (MIMIC)-IV v2.2 dataset.

**Participants:** We randomly selected 100 hospitalizations of adult survivors of critical illness. Four contemporary LLMs (Open AI GPT-4o, o3- and o4-mini, and DeepSeek-R1) and 7 physicians provided independent prognostic estimates for each case (1,100 total estimates; 400 LLM and 700 physician).

**Main outcomes and measures:** For each case, LLMs and physicians used the hospital discharge summary and demographics to predict 6-month mortality (yes/no) and provide their reasoning (free text). We assessed prognostic performance using accuracy, sensitivity, and specificity, and used inductive, qualitative content analysis to characterize reasonings.

**Results:** Mean physician accuracy for predicting mortality was 70.1% (95% CI 63.7-76.4%), with sensitivity of 59.7% (95% CI 50.6-68.8%) and specificity of 80.6% (95% CI 71.7-88.2%). The top-performing LLM (OpenAI o4-mini) accuracy was 78.0% (95% CI 70.0-86.0%), with sensitivity of 80.0% (95% CI 67.4-90.2%) and specificity of 76.0% (95% CI 63.3-88.0%). The difference between mean physician and top-performing LLM accuracy was not statistically significant (p = 0.5). Qualitative analysis revealed similar patterns in LLM and physician expressed reasoning, except that physicians regularly and explicitly reported uncertainty while LLMs did not.

**Conclusion and Relevance:** In this study, LLMs and physicians achieved comparable, moderate performance in predicting 6-month mortality after critical illness, with similar patterns in expressed reasoning. Our findings suggest LLMs could be used to support prognostication in clinical practice but also raise safety concerns due to the lack of LLM uncertainty expression.

**KEY POINTS:** *Question:* How does large language model (LLM) prognostic accuracy and reasoning compare to physicians when predicting 6-month mortality for adult survivors of critical illness?

*Findings:* In this embedded mixed methods study, physicians and large language models had comparable, moderate prognostic accuracy with similar expressed reasoning patterns except that LLMs did not explicitly express uncertainty.

*Meaning:* Large language models may be able to support physician prognostication, although the inability of LLMs to express uncertainty poses an important safety consideration.

## INTRODUCTION

Prognostication is an essential clinical task, as physicians routinely formulate and use prognostic estimates to inform treatment plans and engage in shared decision making.^1,2^ This task is especially important in the context of critical illness, with high-stakes decisions and challenging tradeoffs between life-sustaining therapies, prolonged periods of recovery, and end-of-life care.^2-4^ Furthermore, most patients and families desire prognostic estimates from physicians,^5,6^ including disclosure of uncertainty about likely outcomes.^7^

Despite the importance of prognostication, this task has well-described challenges.^8-10^ For example, studies evaluating physician and nurse prognostication for patients in the intensive care unit (ICU) demonstrate only moderate accuracy and reveal frequent discordance between different clinicians prognosticating for the same patient.^11-14^ Many physicians report high levels of uncertainty when asked to formulate prognostic estimates,^15,16^ which contributes to their hesitation in providing estimates to patients and families.^17^

Multiple scoring systems and machine learning models have been developed to predict likelihood of short- and long-term mortality among critically ill patients.^18-20^ Yet, the use of these models to inform an individual patient’s care remains contested,^21-23^ primarily because they produce population-level estimates that can be challenging to apply to individual patients. Thus, most physicians continue to rely on clinical experience to prognosticate for individual patients.^24-27^

More recently, there is interest in the potential of generative artificial intelligence, specifically large language models (LLMs) with reasoning capacity, to support physicians’ prediction tasks and prognostication efforts for individual patients.^28-30^ The utility of LLMs has been demonstrated in summarizing complex clinical care, categorizing illness severity, and predicting outcomes after specific treatments or surgery.^28-34^ However, it remains unknown how LLMs compare with physician prognostication of mortality, specifically in the heterogenous population of critically ill adults. Given the high-stakes nature of this task and ongoing concerns about LLM biases, “black box” lack of transparency, and halllucinations,^35-41^ it is important not only to evaluate how well LLMs prognosticate, but also to understand how LLMs formulate estimates and express their reasoning.

Therefore, the overarching objective of this mixed methods study was to compare the prognostic performance and expressed reasoning between practicing physicians and contemporary LLMs when predicting 6-month mortality for survivors of critical illness.

## METHODS

### Overall mixed methods study design

We conducted an embedded mixed methods study^42^ using a subset of hospitalized patient cases from the publicly available, deidentified Medical Information Mart for Intensive Care (MIMIC)-IV v2.2 dataset.^43-45^ We elicited prognostic estimates about patients’ 6-month mortality from 4 contemporary LLMs and 7 physicians, including free-text expressed reasoning for each estimate. We quantitatively compared the prognostic performance of LLMs versus that of physicians and qualitatively analyzed and compared the stated reasonings. The study was reviewed and determined not to constitute human subjects research by the University of Wisconsin Institutional Review Board; all physician reviewers complied with the PhysioNet Data Usage Agreement and regulatory requirements.

### Patient Case Selection

The MIMIC-IV v2.2 dataset includes electronic health records from all adult (age ≥ 18) patients admitted to the emergency department (ED) or ICU at a single academic health center in the Northeastern US from 2008 through 2019.^43-45^ MIMIC-IV v2.2 includes date of death linked from state vital records. Given our focus on 6-month mortality after hospitalization, we only included patients who survived to hospital discharge and were discharged to a location other than a hospice facility. We excluded ED-only encounters and hospitalizations without discharge summaries. Among 56,756 hospitalizations for 44,010 unique patients meeting these criteria in the MIMIC-IV v2.2 dataset, we selected a simple random sample of 100 cases. Because our purpose was prognostic performance evaluation and not risk prediction, we balanced the sampled to ensure an equal number of cases who died before and after 6 months and a time-to-death distribution matching the overall cohort.

The adequacy of a 100-case sample size was determined using the R package *KappaSize*, which implements the approach described by Rotondi et al.^46^ Interobserver agreement was assessed for the binary outcome of 6-month survival. With a target Cohen’s κ of 0.70, consistent with rater training benchmarks, and a precision of 0.10 to yield a 95% confidence interval lower bound of 0.60 (moderate agreement), the minimum required sample size was 73 cases per rater.

### Participating Physicians and Prognostication Tasks

We recruited 7 physicians from specialties that routinely prognosticate for critically ill patients (critical care, emergency medicine, internal medicine) to provide prognostic estimates. Based on physician characteristics associated with prognostic accuracy,^47^ we purposefully sampled physicians to include those who had completed (n=4) and were in training (n=3; i.e., residency or fellowship) and to ensure representation of female (n=3) and male (n=4) physicians. We measured physicians’ tolerance of uncertainty using the Physician Intolerance of Uncertainty Scale,^16,48^ with 2 reporting medium and 5 reporting low tolerance.

Physicians received each patient’s hospital discharge summary, age, race/ethnicity, hospital length of stay (LOS), ICU LOS, and discharge disposition. Physicians used this information to answer the following prompts: (1) Do you think the patient will be alive in 6 months? [Yes/No]; (2) What is your level of confidence in your answers? [scale of 1 to 100]; and (3) What is your reasoning for your answers, in 4 sentences or less? [free text response box]. All 7 physicians provided independent prognostic estimates for the 100 hospitalizations, generating 700 total estimates. Physician estimates were collected using a Research Electronic Data Capture (REDCap)^49,50^ data collection form, with physician identity anonymized for analysis.

### Large Language Models and Prognostication Tasks

Four contemporary, high-performing LLMs were selected to provide prognostic estimates for the same 100 hospitalizations: GPT-4o (Azure API version 2024-05-01-preview), o3-mini (2024-12-o1-preview), o4-mini (2025-04-01-preview), and DeepSeek-R1 (2024-05-01-preview). DeepSeek-R1 is an open reasoning model with 685 billion parameters, while all other models are from OpenAI, including reasoning and non-reasoning models. All LLM experiments were performed via the Application Programming Interface (API) in a HIPAA-secure Microsoft Azure cloud environment at the UW-Madison School of Medicine. No patient data were transmitted, stored, or used by OpenAI for training or human review. Interactions with closed-source LLMs were fully HIPAA-compliant, safeguarding patient confidentiality.

LLMs and physicians received the same information about each case and were prompted to perform the same prognostication tasks. Prompts were formatted to ensure compatibility with each model’s training schema and consisted of fixed textual components: a system prompt and a task prompt with responses in JavaScript Object Notation output for analysis. System prompts included general instructions to the model (i.e., “You are an experienced critical care physician with expertise in accurately predicting patient outcomes based on clinical and demographic data.”) Task prompts mirrored the instructions given to physicians, but LLMs were not prompted to report confidence as LLM verbalized confidence is not a reliable measure of risk uncertainty.^51^ Full prompt details are provided in the Supplemental Appendix.

### Quantitative Analysis

We evaluated the prognostic accuracy of physicians and LLMs for predicting 6-month mortality using overall accuracy, sensitivity, and specificity with bootstrapped 95% confidence intervals (CI). Given the balanced dataset, we selected accuracy for the primary comparison. To facilitate comparison with other LLM evaluations, we also calculated F1 scores (a performance metric representing the harmonic mean between precision [positive predictive value] and recall [sensitivity]).^52^ We calculated metrics at aggregated (i.e., pooled physician estimates versus pooled LLM estimates) and individual levels (i.e., individual physicians versus individual models). To best simulate how LLMs might be applied in clinical practice to support physicians, our primary comparison was the pooled physician estimates compared to the top-performing LLM. The primary comparison was assessed using a bootstrap resampling approach with 1,000 iterations. In each iteration, cases were sampled with replacement, and the accuracy difference was computed. Statistical significance was set at p < 0.05. Fleiss’ Kappa scores were also calculated to determine inter-rater agreement between physicians and LLMs. To evaluate differences in performance at varying physician confidence levels, cases were divided into three groups according to the aggregate physician confidence ratings (high confidence, medium confidence, and low confidence) and compared performance across the respective groups.

### Qualitative Analysis

We used inductive qualitative content analysis^53^ to analyze responses to the prompt: “What is your reasoning for your answers?” A team of 4 coders from diverse backgrounds (medicine, critical care, biology) independently annotated reasoning responses for the first 5 patient cases, inductively generating codes representing common patterns. The group met to review, compare, and refine the codes to generate a codebook, which was then independently applied by 3 coders to the next 5 cases. After achieving and confirming inter-coder reliability through consensus review of these initial 110 responses, the remaining cases were divided amongst the 3 coders and coded independently. The team met regularly to review codes, adjudicate questions, and iteratively refine the codebook. After the first round of coding, two coders pursued a directed analysis to identify and characterize hallucinations, specifically comparing LLM and physician responses to the patient information. Investigators then met to conduct a higher-level analysis, identifying patterns and relationships within and among the codes and developing higher-level themes. Divergent cases were sought out to test and refine the developing themes.

## RESULTS

Among included cases, median patient age was 70 years (IQR 58-79), and median Sequential Organ Failure Assessment (SOFA) score at ICU admission was 4.0 (IQR 2.0-5.8). The most frequent discharge dispositions were home (47%) and skilled nursing facility (30%; **Table 1**).

**Table 1.** Patient characteristics among the 100-hospitalization sample.

| Patient Characteristic | N = 100 |
| --- | --- |
| Age in years — median (IQR) | 70 (58-79) |
| Sex – n (%) |  |
| Female | 48 (48%) |
| Male | 52 (52%) |
| Race/Ethnicity — n (%) |  |
| White<sup>a</sup> | 67 (67%) |
| Black/African American<sup>b</sup> | 16 (16%) |
| Hispanic/Latino<sup>c</sup> | 5 (5%) |
| Other<sup>d</sup> | 12 (12%) |
| Maximum SOFA Score<sup>e</sup> — median (IQR) | 4.0 (2.0 to 5.8) |
| ICU Length of Stay — median days (IQR) | 2.1 (3.1) |
| Hospital Length of Stay — median days (IQR) | 6.8 (7.5) |
| Discharge Destination — n (%) |  |
| Home Health Care | 30 (30%) |
| Skilled Nursing Facility | 30 (30%) |
| Home | 17 (17%) |
| Long Term Acute Care Hospital | 12 (12%) |
| Rehab | 5 (5%) |
| Other Acute Care Hospital | 3 (3%) |
| Unknown | 3 (3%) |

a: Includes those who identify as “other European”, “Russian”, and “Brazilian”
b: Includes those who identify as “Cape Verdean”
c: Includes those who identify as “Puerto Rican”, “Dominican”, “Guatemalan”
d: Includes the categories “Unable to obtain”, “Unknown”, “Other”, and “Patient declined to answer”
e: Reported as the maximum SOFA score obtained during the first 24 hours of ICU admission. 2 out of 100 cases excluded from this variable due to missing SOFA scores in the dataset.
Abbreviations: ICU = intensive care unit; IQR = interquartile range; SOFA = sequential organ failure assessment

### Overall performance

Mean, pooled physician accuracy for predicting mortality was 70.1% (95% CI 63.7-76.4%), with sensitivity of 59.7% (95% CI 50.6-68.8%) and specificity of 80.6% (95% CI 71.7%-88.2%). The mean physician F1 score was 66.2% (95% CI 57.3-73.5%). For the top-performing LLM (OpenAI o4-mini), overall accuracy was 78.0% (95% CI 70.0-86.0%), with sensitivity of 80.0% (95% CI 67.4-90.2%), specificity of 76.0% (95% CI 63.3-88.0%), and F1 score of 78.4% (95% CI 68.2-86.6%). Mean, pooled LLM accuracy was 75.0% (95% CI 68.8%-81.3%), with a sensitivity of 78.5% (95% CI 70.3-85.9%), specificity of 71.5% (95% CI 61.5-81.2%), and mean F1 score of 75.1% (95% CI 67.0%-82.3%). Individual physician accuracies ranged from 63.0% to 77.0%; model accuracies ranged from 70.0% to 78.0% (**Figure 1**). The difference between mean pooled physician accuracy and the top-performing LLM was not statistically significant (p =0.5). Inter-rater agreement was moderate among physicians (kappa = 0.52 [95% CI 0.43-0.61]) and LLMs (kappa = 0.59 [95% CI 0.48-0.69]).

**Figure 1.**
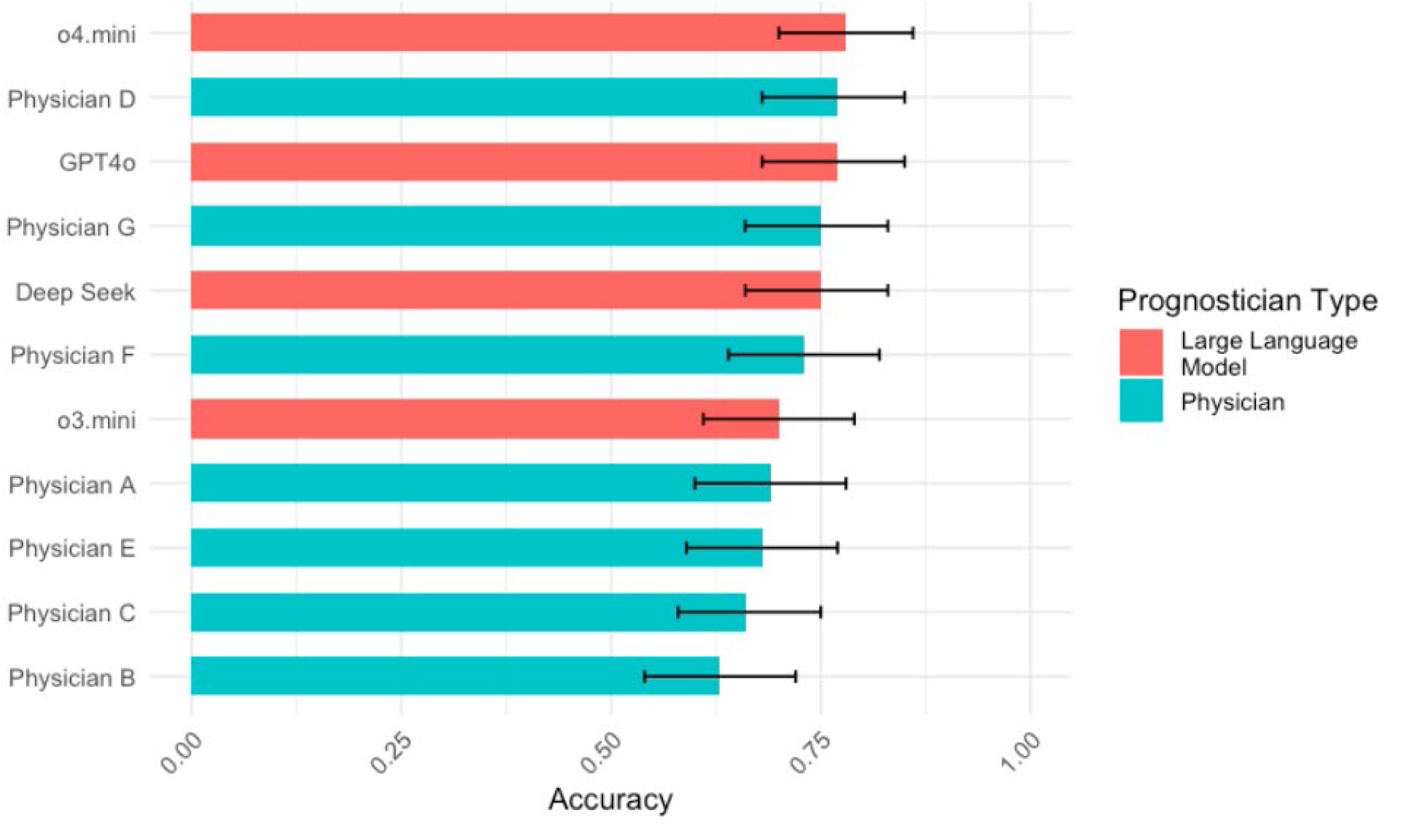
Comparing the overall accuracy of 7 physicians and 4 large language models in estimating 6-month mortality for patients who survived critical illness. Bars represent the overall accuracy for each of the 7 physicians and 4 large language models included in the study, with bootstrapped 95% confidence intervals. The primary comparison (top-performing LLM accuracy versus pooled physician accuracy) was not statistically significant (p=0.50).

### Performance stratified by physician confidence

We categorized cases into low- (lowest quartile of physician confidence), medium- (second and third quartiles), and high-confidence (highest quartile). **Figure 2** illustrates physician and LLM accuracies when stratified by confidence. For cases in the high-confidence group, mean physician accuracy increased to 90.5% (95% CI 85.0%-95.2%) while the top-performing LLM (o4-mini) accuracy was 96.0% (95% CI [88.0%-100.0%]; p= 0.615). In the low-confidence group, mean physician accuracy dropped to 53.2% (95% CI 45.8 -60.2%) while the top-performing LLM (o4-mini) accuracy was 57.9% (95% CI [36.8-78.9%]; p=0.507).

**Figure 2.**
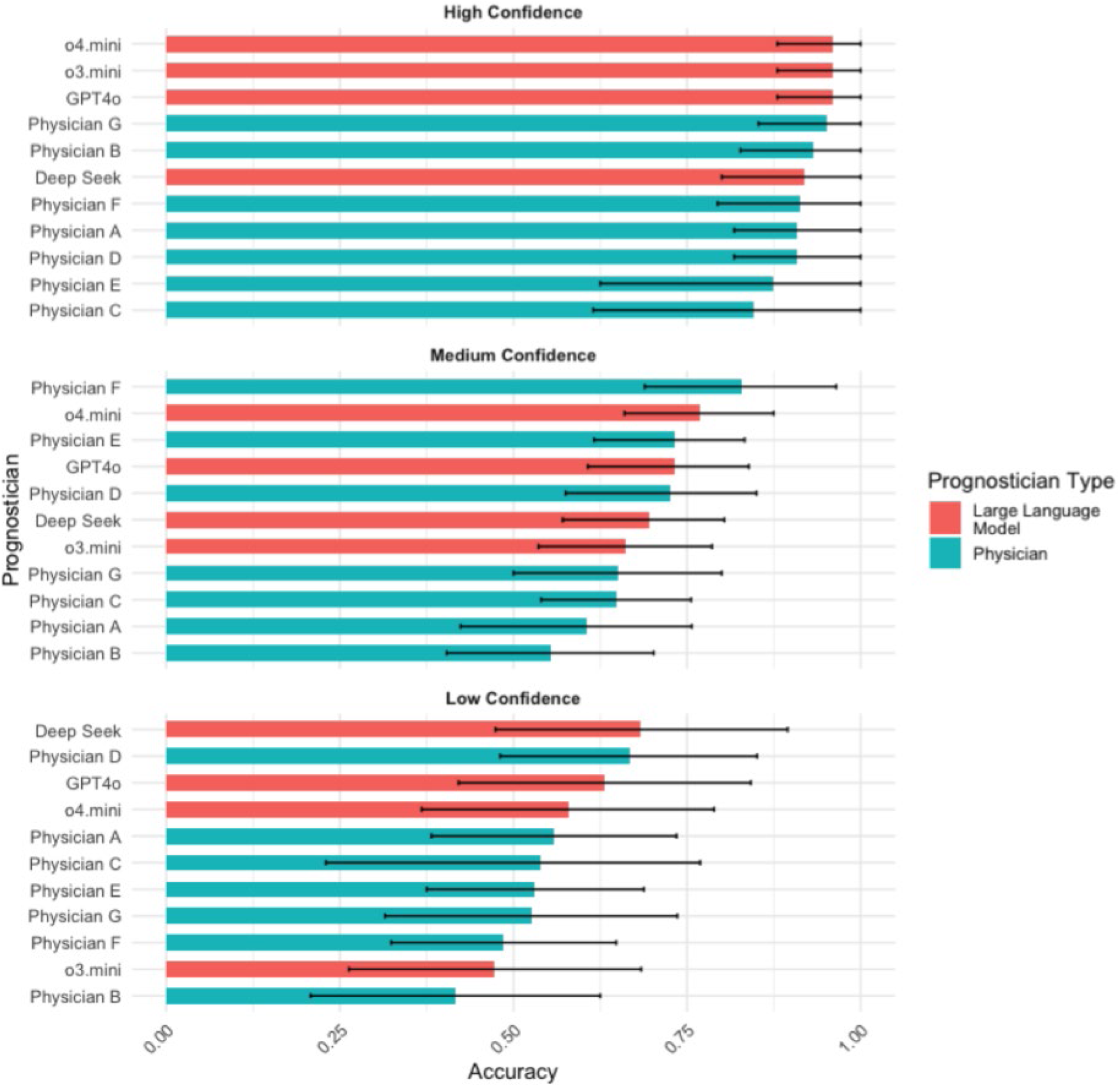
Comparing accuracy of physicians and large language models in predicting 6-month mortality when cases are stratified by physicians’ level of confidence in prediction. Bars represent the overall accuracy for each of the 7 physicians and 4 large language models included in the study, with bootstrapped 95% confidence intervals. Comparisons of the top-performing LLM accuracy with pooled physician accuracy in each group were not statistically significant (low confidence group p= 0.51, medium confidence group p=0.49, high confidence group p=0.62).

### Characterizing stated prognostic reasoning

In our qualitative analysis of responses to the prompt, “What is your reasoning for your answers?,” we identified similarity between LLM and physician free-text narrative comments, in both style and reported patient factors influencing their prognostic estimates. These factors included traditional, static risk factors for mortality such as age, number or extent of comorbidities, functional status, and primary reason for critical illness. For example, one LLM responded, “[A patient in their 80s] with symptomatic severe aortic stenosis (syncope on exertion) left untreated at discharge, plus dementia, CAD, and diabetes, his 6-month mortality risk exceeds 50%…" For the same case, a physician wrote: “The contributing factors to my estimation that the patient will not be alive are his age [(above 80)], underlying dementia, multiple cardiac conditions preceding this hospital stay, and the severe aortic stenosis.” Beyond traditional, static factors, we found physicians and LLMs routinely incorporate more dynamic or time-based factors into their prognostic estimates (**Table 2**). These dynamic features include patient response to medical interventions over time, duration of life-sustaining treatment, and/or duration of ICU or hospital stay.

**Table 2.** Qualitative themes that illustrate how a patient’s dynamic clinical trajectory informed prognostic estimates.

| Major theme | Exemplary quotes |  |
| --- | --- | --- |
|  | Physician | Large language model |
| **Response to medical intervention** | “Significant acute illness w/ longer hospital stay [...], but the underlying problem has been fixed and seems to be responding appropriate to treatment.” (Physician D, Case 7)<br>“The patient is likely to die in the next 6 months because of her severe cardiovascular disease and her tenuous volume status that makes hemodialysis very challenging for her. I don’t see a pathway towards improvement for her, and it is likely she will have recurrent complications and ultimately die.” (Physician F, Case 36) | “Despite the patient’s advanced age and multiple comorbidities [...], she showed clinical improvement during her hospital stay, including stabilization of acute respiratory failure and diuretic management of her pleural effusion” (OpenAI GPT-4o, Case 15)<br>“her hospital course emphasized progressive respiratory compromise despite treatment modifications. These objective findings suggest that survival at 6 months is unlikely.” (OpenAI o3-mini, Case 37) |
| **Duration of life-sustaining treatments** | “No severe complications and patient was quickly extubated and weaned off pressors.” (Physician E, Case 18)<br>“His overall prognosis is poor as he is trached, ventilator dependent as well as G-tube dependent with low likelihood of coming off the ventilator.” (Physician A, Case 6) | “early extubation, rapid wean from IABP and inotropes, and full ambulation by discharge, she demonstrated strong post-operative recovery.” (OpenAI o4-mini, Case 59)<br>“[Patient is in their 80s] with prolonged ICU/ventilator dependence, tracheostomy and PEG placement, [...] her 6-month post-ICU survival odds are poor.” (OpenAI o4-mini, Case 84) |
| **Duration of hospital or ICU stay** | “Reasonably high risk for 6mo mortality [...] however, this was a brief illness that responded well to treatment w/ short hospital stay. [...]” (Physician D, Case 0)<br>“Prolonged ICU stay. High risk for further complications.” (Physician B, Case 38) | “The patient, despite being [in their 80s] with multiple comorbidities [...] demonstrated a relatively rapid recovery from septic shock with stabilization and improvement over a short ICU and hospital stay” (OpenAI o3-mini, Case 0)<br>“Prolonged hospitalization (17.9 days) and recurrent ICU admissions for |
|  |  | hypoxemia suggest advanced disease [...].” (DeepSeek-R1, Case 37) |

While physicians regularly included explicit statements of uncertainty in their responses (“his overall trajectory seems uncertain”), LLMs never explicitly reported being uncertain. However, both LLMs and physicians demonstrated a deliberation process in their responses by weighing favorable and unfavorable factors. For example, “Although advanced age and underlying conditions increase long-term risks, the objective data indicate he was sufficiently robust at discharge, making it likely he will be alive 6 months after ICU discharge” (OpenAI o3-mini).

### “Hallucinations” in LLMs’ expressed reasoning

We identified very few confabulations or factual inaccuracies in LLM responses. These inaccuracies were minor, such as describing a patient as receiving oxygen therapy when only a mention of low oxygen saturation was provided in the discharge summary or misclassifying a beta-blocker medication as anti-coagulation/anti-platelet therapy (**Table 3**). More often, we identified inferences and judgments within LLM responses that were not fully justified based on the information provided. For example, in one response, an LLM describes a patient’s “family support" after discharge as a favorable prognostic factor, while the discharge summary only described family as present at the patient’s bedside. Notably, we also saw this type of not-fully-justified inference in physician responses. For example, a physician described a patient as having “decompensated cirrhosis” when the discharge summary only mentioned chronic liver disease without explicit mention or features of decompensation.

**Table 3.**
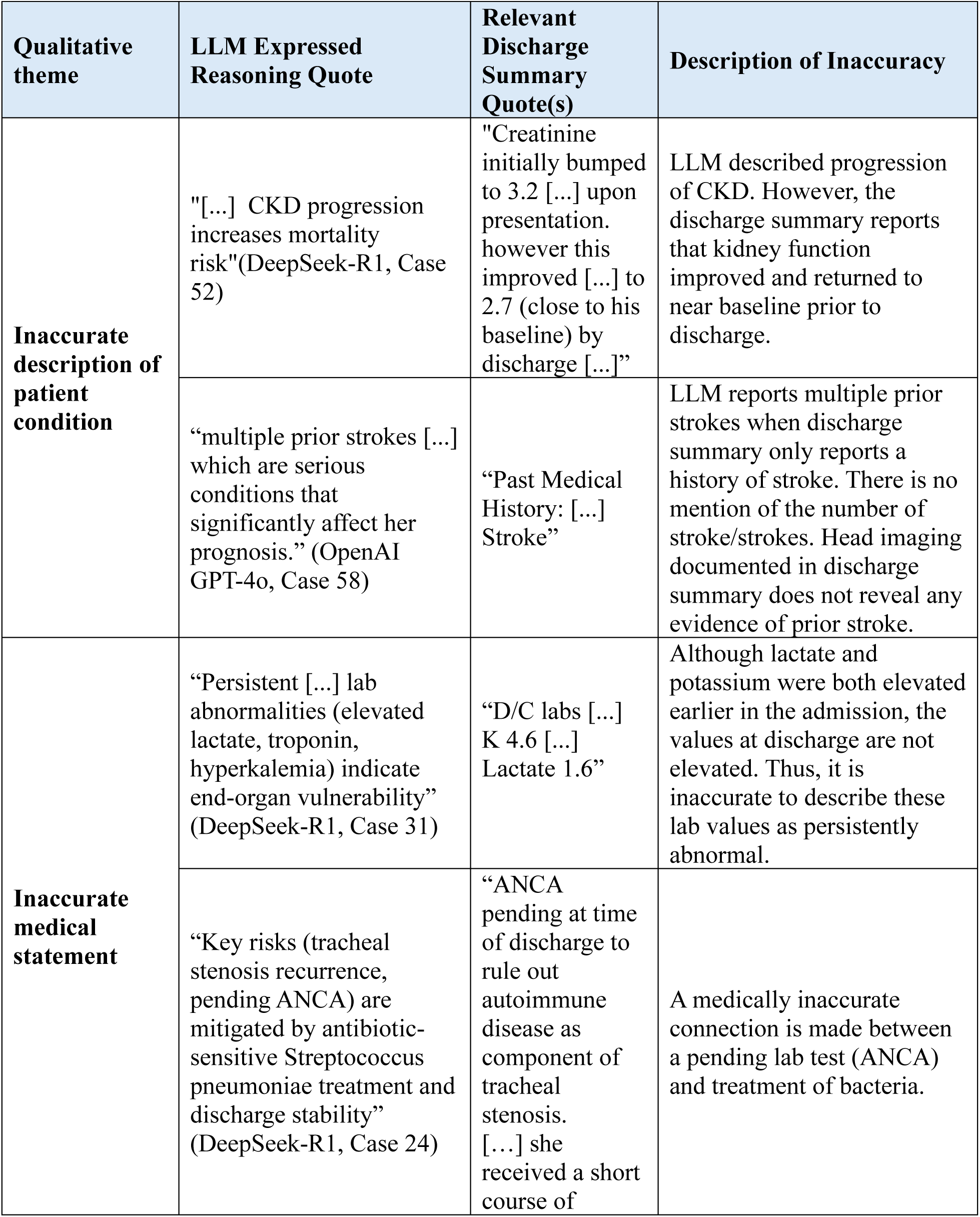
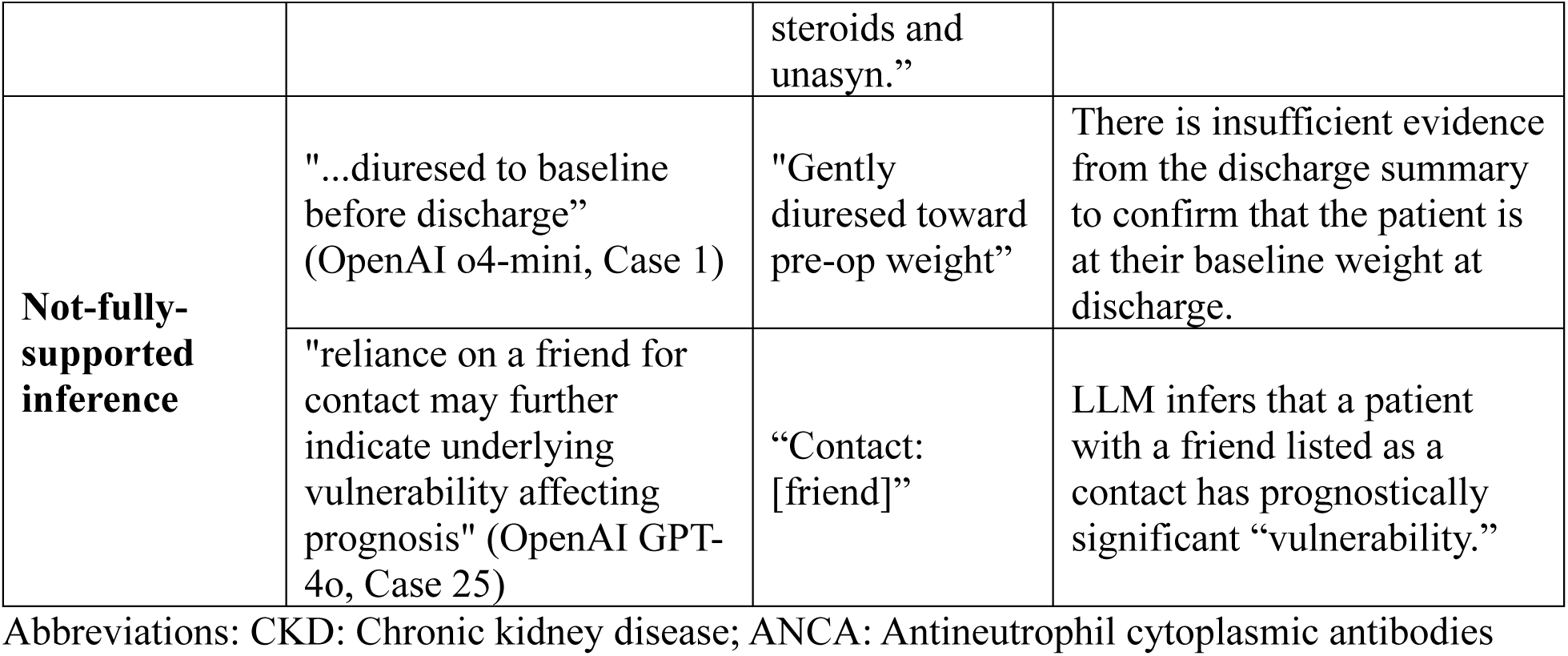
Qualitative themes illustrating inaccuracies identified within LLM expressed reasoning.

| Qualitative theme | LLM Expressed Reasoning Quote | Relevant Discharge Summary Quote(s) | Description of Inaccuracy |
| --- | --- | --- | --- |
| **Inaccurate description of patient condition** | "[...] CKD progression increases mortality risk"(DeepSeek-R1, Case 52) | "Creatinine initially bumped to 3.2 [...] upon presentation. however this improved [...] to 2.7 (close to his baseline) by discharge [...]" | LLM described progression of CKD. However, the discharge summary reports that kidney function improved and returned to near baseline prior to discharge. |
|  | “multiple prior strokes [...] which are serious conditions that significantly affect her prognosis.” (OpenAI GPT-4o, Case 58) | “Past Medical History: [...] Stroke” | LLM reports multiple prior strokes when discharge summary only reports a history of stroke. There is no mention of the number of stroke/strokes. Head imaging documented in discharge summary does not reveal any evidence of prior stroke. |
| **Inaccurate medical statement** | “Persistent [...] lab abnormalities (elevated lactate, troponin, hyperkalemia) indicate end-organ vulnerability” (DeepSeek-R1, Case 31) | “D/C labs [...] K 4.6 [...] Lactate 1.6” | Although lactate and potassium were both elevated earlier in the admission, the values at discharge are not elevated. Thus, it is inaccurate to describe these lab values as persistently abnormal. |
|  | “Key risks (tracheal stenosis recurrence, pending ANCA) are mitigated by antibiotic-sensitive Streptococcus pneumoniae treatment and discharge stability” (DeepSeek-R1, Case 24) | “ANCA pending at time of discharge to rule out autoimmune disease as component of tracheal stenosis. [...] she received a short course of steroids and unasyn.” | A medically inaccurate connection is made between a pending lab test (ANCA) and treatment of bacteria. |
| **Not-fully-supported inference** | "...diuresed to baseline before discharge” (OpenAI o4-mini, Case 1) | "Gently diuresed toward pre-op weight” | There is insufficient evidence from the discharge summary to confirm that the patient is at their baseline weight at discharge. |
|  | "reliance on a friend for contact may further indicate underlying vulnerability affecting prognosis" (OpenAI GPT-4o, Case 25) | “Contact: [friend]” | LLM infers that a patient with a friend listed as a contact has prognostically significant “vulnerability.” |

Abbreviations: CKD: Chronic kidney disease; ANCA: Antineutrophil cytoplasmic antibodies

## DISCUSSION

In this study, we found that physicians and contemporary, high-performing LLMs have moderate and comparable accuracy in predicting 6-month mortality for survivors of critical illness. The accuracy of both physicians and LLMs was very high (>90% correct) when physicians had high confidence in their responses compared to lower-confidence responses (<60% correct). When characterizing how LLMs and physicians express their prognostic reasoning, we found similar patterns. These findings suggest that contemporary LLMs can approximate the prognostic accuracy and emulate the clinical reasoning of practicing physicians.

Our findings have important implications for those considering how best to incorporate LLMs into clinical practice. While the top-performing LLM (OpenAI o4-mini) achieved higher accuracy than the top-performing physician and the pooled physicians, the comparison was not statistically significant, preventing definitive claims about superior performance. We did find variation in the prognostic performance between individual physicians with only moderate inter-rater reliability. This finding suggests that, while LLMs do not surpass top-performing physicians, one potential application for LLMs in clinical practice is to reduce variation in physician-level performance.

Our findings align with prior work examining physician prognostication, including a study by Detsky et al evaluating accuracy in predicting 6-month mortality among critically ill patients. We found similar, moderate prognostic accuracy among physicians in our study and replicated their finding of improved and high prognostic accuracy (∼90%) when physician-reported confidence was high. Our findings add to this work by demonstrating high-performing LLMs also achieve moderate accuracy when performing this task. Together, these findings add to a body of literature that underscores the persistent challenge in achieving high levels of accuracy in predicting longer-term outcomes (e.g., 6-month mortality) for critically ill patients.^12-14^

One potential explanation for this persistent challenge is the irreducible, or aleatoric, uncertainty^54^ inherent to a patient’s future that cannot be minimized or overcome with greater clinical experience, more data, or better models. Indeed, physicians in this study regularly acknowledged this uncertainty when describing how they arrived at prognostic estimates.

Notably, none of the LLM expressed reasonings included an explicit acknowledgement of uncertainty. This may be due to a post-training artifact for LLMs, as annotators may favor responses with more confident language,^55^ or known LLM limitations in producing verbalized confidence levels.^51^ If LLMs are unable or trained not to express uncertainty when uncertainty exists, model output used in clinical practice could convey undue confidence in a particular prediction. This undue confidence could inaccurately influence patient care decisions, which is an important potential safety concern for LLMs being deployed as decision support tools. Taken together, these findings highlight the ongoing need for interventions and tools that accurately identify and manage prognostic uncertainty,^56-58^ rather than focusing primarily on predictive models to overcome it.

In our qualitative analysis, we found similar patterns of expressed reasoning between LLMs and physicians, with both groups drawing on similar, traditional risk factors when prognosticating. More notably, they consider the complex interaction of traditional risk factors with the dynamic features of a patient’s longitudinal clinical course. This finding aligns with recent work incorporating the dynamic clinical trajectory of a patient into prognostic models for short-term hospital outcomes.^59,60^ Our work confirms and advances these efforts, suggesting longitudinal clinical trajectories carry prognostic weight 6 months after critical illness. Modeling efforts should continue to focus on incorporating these dynamic and longitudinal features. Importantly, given the ongoing concerns about LLM “hallucinations,”^38-41^ we found only rare and likely inconsequential instances of LLM factual inaccuracy and no instances of nonsensical reasoning. Both LLMs and physicians made inferences that were plausible but not fully supported by the provided data, suggesting that drawing inferences may be an essential element of prognostication. In prior studies evaluating hallucinations in diagnosis and text summarization, hallucinations are similarly seen among both physicians and LLMs and are often introduced as evidence to support their reasoning.^61,62^

There are several limitations to this study. Physicians and LLMs estimated prognosis using discharge summary notes, as discharge summaries provide a concise narrative of a patient’s longitudinal course. Thus, our findings are reflective of the discharge timepoint and may not reflect prognostic accuracy at other clinically relevant moments. While we elicited a large number of physician estimates, the 7 physicians may not be representative of all physicians. We generated a single LLM estimate per case using one fixed system prompt and a deterministic temperature setting. This design reduces variability and simplifies comparison but does not capture the full range of responses the model might produce under different conditions. Finally, we compared physicians against LLMs but did not evaluate the prognostic performance of physicians supported by LLMs, which is a clinically relevant scenario and should be evaluated in future work.

In conclusion, we found that LLMs and physicians achieved comparable, moderate performance in predicting 6-month mortality after critical illness and reported remarkably similar patterns of reasoning. Our findings suggest LLMs could be used to support prognostication in clinical practice but also raise potential safety concerns given the inability of LLMs to explicitly express uncertainty.

## Data Availability

All data produced in the present study are available upon reasonable request to the authors

## Notes

### Competing Interest Statement

The authors have declared no competing interest.

### Funding Statement

This study did not receive any funding

### Author Declarations

This study was reviewed and determined not to constitute human subjects research by the University of Wisconsin Institutional Review Board. Additonally, all physician reviewers complied with the PhysioNet Data Usage Agreement and regulatory requirements.

